# Attitudes toward COVID-19 pandemic among fully vaccinated individuals: evidence from Greece two years after the pandemic

**DOI:** 10.1101/2022.07.16.22277705

**Authors:** Petros Galanis, Irene Vraka, Aglaia Katsiroumpa, Olga Siskou, Olympia Konstantakopoulou, Theodoros Katsoulas, Theodoros Mariolis-Sapsakos, Daphne Kaitelidou

## Abstract

**Background:** Considering the major effects of COVID-19 pandemic on health, social, economic, and political dimensions of all countries, positive attitudes toward COVID-19 pandemic are essential to control the pandemic. In our study, we investigated attitudes toward COVID-19 pandemic among fully COVID-19 vaccinated individuals two years after the pandemic and we identified predictors of attitudes.

**Materials and Methods:** We conducted an on-line cross-sectional study with 815 fully COVID-19 vaccinated individuals in Greece during May 2022. A self-administered and valid questionnaire was disseminated through social media platforms. We measured socio-demographic variables and COVID-19-related variables as potential predictors of attitudes toward COVID-19 pandemic. The outcome variable was attitudes toward COVID-19 pandemic (compliance with hygiene measures, trust in COVID-19 vaccination, fear of COVID-19, and information regarding the COVID-19 pandemic and vaccination).

**Results:** We found a very high level of compliance with hygiene measures, a high level of trust and information about the COVID-19 pandemic and vaccination, and a moderate level of fear of COVID-19. Also, we identified that females, participants with a higher educational level, those with a chronic disease, those with a better self-perceived physical health, and those without a previous COVID-19 diagnosis adhered more in hygiene measures. Trust in COVID-19 vaccination was higher among females, older participants, those with a higher educational level, those with a better self-perceived physical health, and those without a previous COVID-19 diagnosis. Moreover, females, older participants, those with a higher educational level, those with a chronic disease, those with a better self-perceived physical health, those that received a flu vaccine in previous season, and those without a previous COVID-19 diagnosis experienced more fear of the COVID-19. Finally, level of information regarding COVID-19 pandemic and vaccination was higher for participants with a higher educational level, those without a chronic disease, those with a better self-perceived physical health, and those that received a flu vaccine in previous season.

**Conclusions:** Understanding predictors of attitudes toward COVID-19 pandemic among fully vaccinated individuals is crucial for developing appropriate public health campaigns in the future. Vaccination should be accompanied by positive attitudes in order to decrease the frequency of negative outcomes of COVID-19, such as hospitalization, complications and mortality.

## Introduction

COVID-19 pandemic is evolving and it is estimated that about 560 million people were infected with COVID-19 pandemic and about 6.3 million people have died until to date (13 July 2022) [1]. Breakthrough infections and new contagious variants of SARS-CoV-2 continue to threaten individuals even those that had completed the COVID-19 vaccine course [2, 3]. Considering the major effects of COVID-19 pandemic on health, social, economic, and political dimensions of all countries, positive attitudes toward COVID-19 pandemic are essential to control the pandemic. Thus, understanding the attitudes of the general population and identifying potential predictors could help policy makers to achieve the outcomes of planed behavior [4, 5]. A systematic review identified 43 studies worldwide that examined the attitudes of the individuals toward COVID-19 and all of them reported a positive attitude [6].

Positive attitudes such as adherence with COVID-19 public health guidelines are influenced by several factors, e.g. older age, female gender, trust in governments, information through traditional news media, and high self-perceived threaten of the COVID-19 [7]. Moreover, COVID-19 vaccination uptake is another important dimension of positive attitudes toward COVID-19 and the available evidence suggests that the vaccination rate is higher among males, white people, older people, those with higher socioeconomic status, those with a chronic condition, and those that are well informed about COVID-19 vaccines [8]. Positive attitudes toward COVID-19 continue to be essential to curb COVID-19 infection rates and there is still interest in investigating potential predictors of these attitudes.

On March 2020, the Greek government announced the first lockdown in Greece due to the first confirmed COVID-19 cases in the country [9]. Two years later, Greece has been among countries with the highest mortality rates from COVID-19 worldwide with 2953 deaths per one million population [1]. Meanwhile, safe and effective COVID-19 vaccines [10, 11] were administrated by early 2021 in Greece. Considering that the rate of completed vaccinations in Greece is high and equal to the rate among countries in the European Union (73.6% vs. 73.3%) [12] the assessment of public attitudes toward COVID-19 pandemic could be critical predicting future attitudes toward COVID-19 vaccination and COVID-19 pandemic in general.

Thus, given the importance of the issue, we investigated attitudes toward COVID-19 pandemic among fully COVID-19 vaccinated individuals in Greece two years after the pandemic and we identified predictors of attitudes.

## Methods

### Study design

We conducted a cross-sectional study in Greece during May 2022. We created an anonymous version of the study questionnaire in Greek using Google forms and we disseminated it through social media platforms. Thus, a convenience sample was obtained. Participation in the study was allowed for individuals aged 18 years or above that understand the Greek language and have completed the COVID-19 vaccine course (primary doses and booster dose). Participation in the study was anonymous and voluntary. Moreover, we followed the principles of the Declaration of Helsinki, while the study protocol was approved by the Ethics Committee of Department of Nursing, National and Kapodistrian University of Athens (reference number; 370, 02-09-2021).

### Predictor and outcome variables

We measured socio-demographic variables and COVID-19-related variables as potential predictors of attitudes toward COVID-19 pandemic. In particular, we measured gender (male or female), age (continuous variable), marital status (single, married, divorced, or widow), educational level (elementary school, high school, or university degree), MSc/PhD degree (yes or no), chronic disease (yes or no), self-assessment of physical health (very poor, poor, moderate, good, or very good), influenza vaccination during 2021 (yes or no), previous COVID-19 diagnosis (yes or no), and COVID-19-related death in family members/friends (yes or no).

The outcome variable was attitudes toward COVID-19 pandemic and was measured with a valid questionnaire [13]. The questionnaire includes 16 items and four factors (compliance with hygiene measures, trust in COVID-19 vaccination, fear of COVID-19, and information regarding the COVID-19 pandemic and vaccination). Internal reliability of the questionnaire in our study was very good since Cronbach’s coefficient alpha was 0.71 for the factor “compliance with hygiene measures”, 0.78 for the factor “trust in COVID-19 vaccination”, 0.85 for the factor “fear of COVID-19”, and 0.81 for the factor “information regarding the COVID-19 pandemic and vaccination”. Responses for the 16 items range from 0 (totally disagree) to 10 (totally agree). Also, a total score from 0 to 10 is calculated for each factor. Higher values indicate higher level of compliance, trust, fear, and information.

### Statistical analysis

We use absolute (n) and relative (%) frequencies to present categorical variables. Continuous variables are presented as mean, standard deviation, median, minimum value, and maximum value. The outcome variables were continuous variables that followed normal distribution. Therefore, we performed univariate and multivariable linear regression analysis to assess the impact of predictor variables on attitudes toward COVID-19 pandemic. We calculated unadjusted and adjusted coefficient beta for each predictor with corresponding 95% confidence intervals (CI) and p-values. A p-value < 0.05 was considered statistically significant. Statistical analysis was performed with the Statistical Package for Social Sciences software (IBM Corp. Released 2012. IBM SPSS Statistics for Windows, Version 21.0. Armonk, NY: IBM Corp.).

## Results

Socio-demographic characteristics and COVID-19-related characteristics of the participants are presented in Table 1. Mean age of 815 participants was 37 years, while 76.1% of them were females, 54% were singles, and 72.4% had a University degree. Moreover, 23.3% of the participants suffered from a chronic disease and 82.4% of them considered their physical health as good/very good. Among the participants, 33.1% received a flu vaccine in previous season, 50.9% have been diagnosed with COVID-19 during the pandemic and 31.3% had family members/friends who had died because of COVID-19.

**Table 1.** Socio-demographic characteristics and COVID-19-related characteristics of the participants.

| Characteristics | N | % |
| --- | --- | --- |
| Gender |  |  |
| Males | 195 | 23.9 |
| Females | 620 | 76.1 |
| Age (years)<sup>a</sup> | 37.0 | 13.3 |
| Marital status |  |  |
| Singles | 440 | 54.0 |
| Married | 290 | 35.6 |
| Divorced | 75 | 9.2 |
| Widowed | 10 | 1.2 |
| Educational level |  |  |
| Elementary school | 0 | 0.0 |
| High school | 225 | 27.6 |
| University degree | 590 | 72.4 |
| MSc/PhD degree |  |  |
| No | 330 | 40.5 |
| Yes | 485 | 59.5 |
| Chronic disease |  |  |
| No | 625 | 76.7 |
| Yes | 190 | 23.3 |
| Self-perceived physical health |  |  |
| Very poor | 0 | 0.0 |
| Poor | 5 | 0.6 |
| Moderate | 90 | 11.0 |
| Good | 400 | 49.1 |
| Very good | 320 | 39.3 |
| Influenza vaccination in previous season |  |  |
| No | 545 | 66.9 |
| Yes | 270 | 33.1 |
| Previous COVID-19 diagnosis |  |  |
| No | 400 | 49.1 |
| Yes | 415 | 50.9 |
| COVID-19-related death in family members/friends |  |  |
| No | 560 | 68.7 |
| Yes | 255 | 31.3 |

<sup>a</sup> mean, standard deviation

Descriptive statistics for the four factors of the questionnaire that we used to measure attitudes toward COVID-19 pandemic are shown in Table 2. Mean value on the factor “compliance with hygiene measures” was 9.0 indicating a very high level of compliance, while mean value on the factor “trust in COVID-19 vaccination” (7.0) denoted a high level of trust. Participants’ fear of the COVID-19 was moderate (mean=5.6), while the level of information about the COVID-19 pandemic and vaccination was high (mean=8.1).

**Table 2.** Descriptive statistics for the four factors that measure attitudes toward COVID-19 pandemic.

| **Factors** | **Mean** | **Standard deviation** | **Median** | **Minimum value** | **Maximum value** |
| --- | --- | --- | --- | --- | --- |
| Compliance with hygiene measures | 9.0 | 1.1 | 9.5 | 4.5 | 10 |
| Trust in COVID-19 vaccination | 7.0 | 1.6 | 7.1 | 2.4 | 9.4 |
| Fear of the COVID-19 | 5.6 | 2.0 | 5.6 | 1.2 | 9.6 |
| Information regarding the COVID-19 pandemic and vaccination | 8.1 | 1.5 | 8.0 | 3.0 | 10.0 |

Univariate and multivariable linear regression analysis with attitudes toward COVID-19 pandemic as the dependent variables are shown in Table 3 and 4. We found that females, married participants, those with a higher educational level, those with a chronic disease, those with a better self-perceived physical health, and those without a previous COVID-19 diagnosis adhered more in hygiene measures. Trust in COVID-19 vaccination was higher among females, older participants, married participants, those with a higher educational level, those with a better self-perceived physical health, those without a previous COVID-19 diagnosis, and those without a COVID-19-related death in family members/friends. Moreover, females, older participants, married participants, those with a higher educational level, those with a chronic disease, those with a better self-perceived physical health, those that received a flu vaccine in previous season, and those without a previous COVID-19 diagnosis experienced more fear of the COVID-19. Finally, level of information regarding COVID-19 pandemic and vaccination was higher for participants with a higher educational level, those without a chronic disease, those with a better self-perceived physical health, those that received a flu vaccine in previous season, and those without a COVID-19-related death in family members/friends.

**Table 3.**
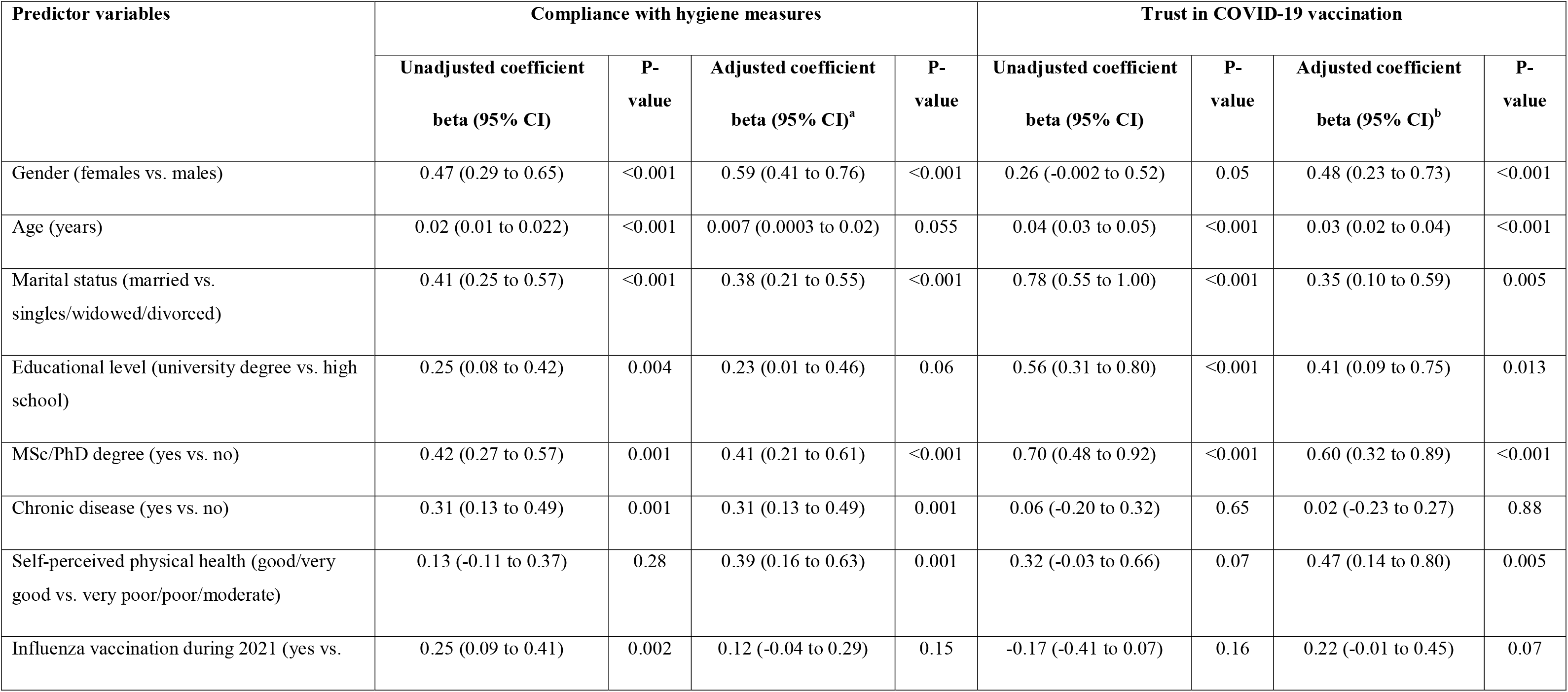
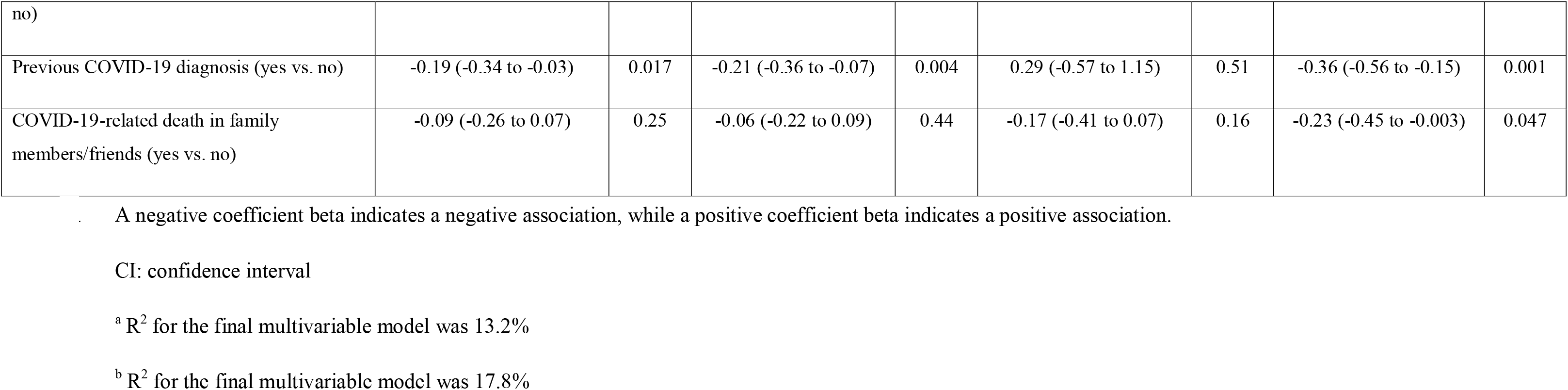
Univariate and multivariable linear regression analysis with individuals’ compliance with hygiene measures and trust in COVID-19 vaccination as the dependent variables.

**Table 4.** Univariate and multivariable linear regression analysis with individuals’ fear of COVID-19 and information regarding COVID-19 pandemic and vaccination as the dependent variables.

| Predictor variables | Fear of COVID-19 |  |  |  | Information regarding COVID-19 pandemic and vaccination |  |  |  |
| --- | --- | --- | --- | --- | --- | --- | --- | --- |
|  | Unadjusted coefficient<br>beta (95% CI) | P-value | Adjusted coefficient<br>beta (95% CI)<sup>a</sup> | P-value | Unadjusted coefficient<br>beta (95% CI) | P-value | Adjusted coefficient<br>beta (95% CI)<sup>b</sup> | P-value |
| Gender (females vs. males) | 0.12 (-0.20 to 0.44) | 0.45 | 0.43 (0.11 to 0.74) | 0.008 | -0.29 (-0.53 to -0.05) | 0.017 | -0.17 (-0.41 to 0.07) | 0.17 |
| Age (years) | 0.04 (0.03 to 0.05) | <0.001 | 0.01 (0.001 to 0.03) | 0.035 | 0.006 (-0.002 to 0.01) | 0.13 | -0.004 (-0.01 to 0.005) | 0.37 |
| Marital status (married vs. singles/widowed/divorced) | 0.74 (0.46 to 1.02) | <0.001 | 0.41 (0.11 to 0.72) | 0.008 | 0.14 (-0.07 to 0.35) | 0.19 | 0.03 (-0.20 to 0.26) | 0.80 |
| Educational level (university degree vs. high school) | 0.93 (0.63 to 1.22) | <0.001 | 0.46 (0.04 to 0.88) | 0.032 | 0.06 (0.17 to 0.28) | 0.63 | 0.45 (0.13 to 0.77) | 0.006 |
| MSc/PhD degree (yes vs. no) | 0.73 (0.46 to 1.00) | <0.001 | 0.19 (-0.17 to 0.55) | 0.30 | 0.22 (0.02 to 0.43) | 0.034 | 0.50 (0.23 to 0.78) | <0.001 |
| Chronic disease (yes vs. no) | 0.39 (0.07 to 0.71) | 0.016 | 0.37 (0.05 to 0.69) | 0.022 | -0.51 (-0.75 to -0.23) | <0.001 | -0.43 (-0.67 to -0.19) | 0.001 |
| Self-perceived physical health (good/very good vs. very poor/poor/moderate) | 0.15 (-0.27 to 0.58) | 0.47 | 0.43 (0.01 to 0.85) | 0.043 | 0.59 (0.27 to 0.90) | <0.001 | 0.42 (0.09 to 0.74) | 0.011 |
| Influenza vaccination during 2021 (yes vs. no) | 0.79 (0.51 to 1.07) | <0.001 | 0.49 (0.20 to 0.79) | 0.001 | 0.67 (0.44 to 0.90) | <0.001 | 0.77 (0.54 to 0.99) | <0.001 |
| Previous COVID-19 diagnosis (yes vs. no) | -0.64 (-0.91 to -0.37) | <0.001 | -0.57 (-0.83 to -0.32) | <0.001 | -0.39 (-0.61 to -0.17) | 0.001 | -0.17 (-0.36 to 0.03) | 0.09 |
| COVID-19-related death in family members/friends (yes vs. no) | 0.10 (-0.19 to 0.39) | 0.50 | 0.02 (-0.27 to 0.30) | 0.91 | -0.20 (-0.42 to 0.02) | 0.08 | -0.24 (-0.46 to -0.03) | 0.028 |

1 A negative coefficient beta indicates a negative association, while a positive coefficient beta indicates a positive association.
CI: confidence interval
<sup>a</sup> R<sup>2</sup> for the final multivariable model was 12.0%
<sup>b</sup> R<sup>2</sup> for the final multivariable model was 10.3%

## Discussion

We conducted a cross-sectional study among fully COVID-19 vaccinated individuals in Greece in order to investigate their attitudes toward COVID-19 pandemic. To the best of our knowledge, this is the first study that examines this issue among fully COVID-19 vaccinated individuals. The study findings are extremely encouraging, since two years after the COVID-19 pandemic our participants expressed a very high level of compliance with hygiene measures and a high level of trust in COVID-19 vaccination. Also, our results shown that the level of information about the COVID-19 pandemic and vaccination was high, while fear of the COVID-19 was moderate. Moreover, we identified several socio-demographic characteristics and COVID-19-related characteristics of the participants as predictors of attitudes toward COVID-19 pandemic.

We found that compliance with hygiene measures was more prevalent in females. Available literature supports this finding [14–17] and could be explained by females’ tendency to practice socially acceptable behavior [18, 19]. This finding is interesting since it is well known that COVID-19 mortality is higher for males compared to females [20, 21]. In our study increased educational level was associated with greater compliance with hygiene measures. This finding is confirmed by studies conducting both in the general population and healthcare workers [22, 23]. Similar studies regarding the swine influenza pandemic in Saudi Arabia and the severe acute respiratory syndrome in Hong Kong confirm the positive effect of education on public attitudes and practices [24, 25]. Higher educational level could help individuals to have a better understanding of the information with regards to the COVID-19 pandemic. Therefore, people in a higher educational level could improve their knowledge and adopt positive preventive practices. Our results support the finding of the literature that chronic conditions are associated with higher level of compliance [26–28]. This finding is not surprising since it is well known that negative outcomes of COVID-19 (e.g. complications, hospitalization, and fatality) are more common among patients with chronic diseases (e.g. diabetes, hypertension, and respiratory chronic diseases) [29, 30]. Therefore, people with morbidities take more precautions in protecting themselves against COVID-19 compared to healthy people.

According to our results, level of trust in COVID-19 vaccination was higher among females, older participants, those with a higher educational level, those without a previous COVID-19 diagnosis, and those with better self-perceived physical health. These findings are confirmed by evidence since older age, higher socioeconomic status, and higher self-perceived COVID-19 vulnerability are the main predictors of COVID-19 vaccination uptake in the general population [8]. In addition, higher educational level and absence of COVID-19 infection have already proven to be independent predictors of COVID-19 vaccination uptake [31, 32]. Additional data from pregnant women reveal that uptake rate is higher among older women and women without a previous COVID-19 diagnosis [33]. Moreover, two recent studies [34, 35] identified female gender as a predictor of individuals’ willingness to accept a first COVID-19 booster dose, while another study [36] found that people with better self-perceived physical health and people without a previous COVID-19 diagnosis had higher odds of accepting a second booster dose.

In our study, females experienced more fear of COVID-19 than males. Several studies both in the general population and healthcare workers confirm that fear of COVID-19 is higher in females [37–41]. In general, females experience the COVID-19 pandemic worse than males, since the prevalence of mood and anxiety disorders are higher in females than males [42, 43]. The relationship between gender and COVID-19 experiences could be attributed to social, biological, and psychological differences between males and females. For instance, females tend to experience more anxiety and psychological distress than males [44]. Moreover, we found that level of fear of COVID-19 was higher among individuals with a chronic disease. This finding echoes the results of similar research which shows a positive relationship between morbidity and fear of COVID-19 [45–47]. Individuals with a chronic condition may be afraid of COVID-19 due to the established association between morbidity and COVID-19-related mortality [29, 30]. In a similar way, we found that increased age was associated with increased level of fear of COVID-19. A systematic review confirms the positive relationship between age and fear of COVID-19 [41]. It is reasonable older individuals to be more afraid of COVID-19 since older age is one of the main predictors of COVID-19-related mortality [48, 49]. In our study, the higher educational level was significantly associated with fear of COVID-19. The impact of education on fear is controversial since two studies [50, 51] in Peru and Iran found a positive relationship between age and fear, while two studies [52, 53] in China and Colombia found the opposite result. Interestingly, a large study with 14,558 participants in 48 countries found that less educated participants experienced increased fear of COVID-19 [54]. Reduced access to health care services and difficulties in understanding health promotion media messages [55] and weaker tendency to respond to health promotion media messages [56] could explain fear of people with lower education.

There is limited evidence regarding the factors that influence information in COVID-19 era since research until now has just focused on the information seeking and the sources of information. We found that level of information regarding COVID-19 pandemic and vaccination was higher for participants with a higher educational level. COVID-19 pandemic affects more vulnerable populations such as less educated with worse health status [57]. A similar study [58] in China found that vulnerable groups (e.g. older, females, and unemployed) expressed more concerns about lack of timely information regarding COVID-19. Therefore, policy makers should provide these vulnerable groups with more adequate and timely information. On the other hand, individuals with a higher educational level have higher expectations for information disclosure since they are more eager to get full information regarding COVID-19 pandemic [58]. Achieving a high level of information is essential to promote preventive behaviors against COVID-19 among the public since there is a direct relation between COVID-19 information and promotion of individual health [59, 60].

### Limitations

Our study had several limitations. First, we used a convenience sample and participants were recruited through social media. Thus, our sample could not be representative of the general population of Greece and we cannot generalize our findings. Studies using nationally representative samples could add more information in this research topic. Also, we used a self-reported questionnaire to collect data that can be influenced by social desirability. Moreover, we investigated only a few socio-demographic characteristics and COVID-19-related characteristics as potential predictors of fear of attitudes toward COVID-19 pandemic. Therefore, future studies should expand research investigating other potential predictors, e.g. psychological variables, personality characteristics, social media variables, information sources, etc. Finally, cross-sectional studies like our study could never establish a causal relation between independent variables and attitudes toward COVID-19 pandemic.

## Conclusions

Understanding predictors of attitudes toward COVID-19 pandemic among fully vaccinated individuals is crucial for developing appropriate public health campaigns in the future. Fully vaccinated people may be feel safe against COVID-19 adopting then less positive attitudes, e.g. less wearing of face masks and non adaptation of physical distancing. In addition, COVID-19-related deaths among fully vaccinated individuals are occurred since no vaccine could have perfect effectiveness. This fact could threaten public trust in COVID-19 vaccination, leading to disappointment and negative attitudes. Therefore, policy makers should always keep in mind fully vaccinated people in order to maintain or even improve positive attitudes toward COVID-19 pandemic in this group. Vaccination against COVID-19 is a tremendous weapon to control the pandemic but it is not the only one. Vaccination should be accompanied by positive attitudes toward COVID-19 pandemic in order to decrease transmission mode of the virus and frequency of negative outcomes of COVID-19, e.g. hospitalization and COVID-19-related deaths.

## Data Availability

All data produced in the present study are available upon reasonable request to the authors

## Acknowledgments

None

